# Do children evaluated for maltreatment have higher subsequent emergency department and inpatient care utilization compared to a general pediatric sample?

**DOI:** 10.1101/2022.06.10.22276264

**Authors:** Yuerong Liu, Megan Shepherd-Banigan, Kelly Evans, Laura Stilwell, Lindsay Terrell, Jillian Hurst, Elizabeth J. Gifford

## Abstract

**Background:** Child maltreatment leads to substantial adverse health outcomes, but little is known about acute health care utilization patterns after children are evaluated for a concern of maltreatment at a child abuse and neglect medical evaluation clinic.

**Objective:** To quantify the association of having a child maltreatment evaluation with subsequent acute health care utilization among children from birth to age three.

**Participants and Setting:** Children who received a maltreatment evaluation (N = 367) at a child abuse and neglect subspecialty clinic in an academic health system in the United States and the general pediatric population (N = 21,231).

**Methods:** We conducted a retrospective cohort study that compared acute health care utilization over 18 months between the two samples using data from electronic health records. Outcomes were time to first emergency department (ED) visit or inpatient hospitalization, maltreatment-related ED use or inpatient hospitalization, and ED use or inpatient hospitalization for ambulatory care sensitive conditions (ACSCs). Multilevel survival analyses were performed.

**Results:** Children who received a maltreatment evaluation had an increased hazard for a subsequent ED visit or inpatient hospitalization (hazard ratio [HR]: 1.3, 95% confidence interval [CI]: 1.1, 1.5) and a maltreatment-related visit (HR: 4.4, 95% CI: 2.3, 8.2) relative to the general pediatric population. A maltreatment evaluation was not associated with a higher hazard of health care use for ACSCs (HR: 1.0, 95% CI: 0.7, 1.3).

**Conclusion:** This work can inform targeted anticipatory guidance to aid high-risk families in preventing future harm or minimizing complications from previous maltreatment.

## Introduction

Child maltreatment is a major public health problem leading to significant pediatric morbidity and mortality (Vaithianathan et al., 2018). In 2019 alone, roughly 2.4 million children in the United States were subject of maltreatment reports and over one-third of maltreatment victims were younger than four years (U.S. Department of Health & Human Services, 2021). Child maltreatment is associated with an elevated risk of common and high-cost pediatric illnesses, which result in a substantial health care burden in terms of both short- and long-term health care use (Peterson et al., 2018). Therefore, an understanding of health care use patterns of potential maltreated children may help prevent escalating or recurrent subsequent maltreatment and reduce inappropriate use of health care.

Children who have been previously maltreated are at high risk of developing diseases, illnesses, or conditions because of new maltreatment (Hindley et al., 2006) or complications from prior maltreatment (Huffhines & Jackson, 2019). These children are likely to experience recurrence of maltreatment (Hindley et al., 2006) and to receive acute health care service for injuries such as fractures and burns (Hunter & Bernstein, 2019; King et al., 2015) that may have resulted from an act related to maltreatment. They may also use acute health care due to increased chance of developing chronic health conditions that may be complicated by previous maltreatment, such as asthma, obesity, diabetes, and eczema (Huffhines & Jackson, 2019; Jackson et al., 2016; Jee et al., 2006; Lanier et al., 2010). This raises important concerns for health care providers in assessing pediatric health outcomes and risk of maltreatment recurrence for children who have a previous concern of maltreatment.

Consistent evidence has shown that compared to children in the general population, children who have experienced maltreatment are high utilizers of emergency department (ED) and inpatient hospitalization (Guenther et al., 2009; Kuang et al., 2018; O’Donnell et al., 2010). Several prevalence studies have estimated incidences of maltreatment-related ED visits or hospitalization among the general pediatric population (Hunter & Bernstein, 2019; King et al., 2015). However, prior studies have focused primarily on patterns of health service use *before* children were diagnosed with maltreatment or involved in child protective service systems (Kuang et al., 2018; O’Donnell et al., 2010). Few studies have examined children’s health care use or maltreatment-related medical encounters after they were identified as being at risk for maltreatment.

With such a high risk of acute health care use and development of undiagnosed or untreated chronic health conditions (e.g., Lanier et al., 2010), children at risk for maltreatment are likely to have preventable ED or hospital admissions (Szilagyi et al., 2015). Some of these chronic conditions (e.g., asthma) are considered to be ambulatory care sensitive conditions (ACSCs) for which an ED visit or hospital admission could be prevented by proper treatment in primary care (Burgdorf & Sundmacher, 2014). Hospitalization and ED visit for ACSCs could serve as an important indicator to quantify health care use disparities among children with previous concerns for maltreatment and children of the general population (Lichtl et al., 2017). Despite this, no research has focused on the prevalence of ACSCs in pediatric patients who have been evaluated for concerns of maltreatment.

Healthcare encounters are one of the only settings in which nearly all young children in the United States are regularly evaluated by a non-caregiver, which can provide potential opportunity to identify and prevent adverse pediatric health outcomes among children at risk of maltreatment. Given the high risk of maltreatment recurrence (Hindley et al., 2006), potential for ongoing health complications related to past maltreatment (Jonson-Reid et al., 2012), and improper use of acute health care for chronic conditions (Szilagyi et al., 2015), it is imperative to assess patterns of high-cost, intensive care use (e.g., ED and inpatient visits) among children with previous concerns of maltreatment. This could help facilitate our understanding of how health care systems could provide sentinel information about these children’s wellbeing over time and ameliorate negative health outcomes.

Evidence as to whether or not potential maltreatment is related to higher rates of subsequent health care use is limited. Moreover, pediatric studies that evaluated health care use of maltreated children have not comprehensively considered all these health care use patterns together. Our study addresses this research gap to improve recommendations for care of this high-risk population and prevent escalating or recurrent maltreatment.

This study used information from the electronic health records (EHR) data for children from birth to three years old who were medically evaluated at a child abuse and neglect subspecialty clinic in a teaching and research hospital in the U.S. The aim of this study was to quantify the association of being evaluated for a concern of maltreatment with subsequent acute health care utilization. We assessed three health care utilization outcomes over 18 months after a child received an evaluation for maltreatment, including (1) ED use and inpatient hospitalization, (2) child maltreatment-related ED use and inpatient hospitalization, and (3) ED use and inpatient hospitalization for ACSCs.

## Methods

### Data Source

Data came from electronic health records (EHR) spanning 3/1/2013 to 6/30/2019 from a large academic health system in the southeastern United States. This health system serves about 85% of children in its primary catchment area. There are one tertiary care and two community-based hospitals, each with an ED. Healthcare is coordinated by utilizing a single EHR system that captures patients’ information from a network of primary care and specialty clinics. The university Institutional Review Board approved this study.

### Study Population

This study used a retrospective cohort design comparing children who received maltreatment evaluation and the general pediatric population. Based on EHR data extracted from the health system, we constructed a cohort of children from birth to three years who (1) resided in a single county that serves as a primary catchment area for the health system, and (2) had either an evaluation for maltreatment at the child abuse and neglect subspecialty clinic or a well-child visit in the health system between 3/1/ 2013 and 12/1/2017. No other exclusion restrictions were applied.

We extracted EHR data for child demographic characteristics, county of residence, health insurance type, date a child was medically evaluated by a child abuse and neglect clinician (applicable to sample evaluated for a concern of maltreatment), date a child received a well-child visit (applicable to the general pediatric population), and date that a child had an ED visit or inpatient hospitalization. ED and inpatient visits were linked to diagnostic (ICD-9-CM or ICD-10-CM) and e-codes to assess whether these visits were related to maltreatment or ACSCs.

### Measures

#### Independent Variable

Our independent variable is *whether or not a child received a medical evaluation for a concern of maltreatment*. Hospitals and health care systems often utilize a specialty clinic to perform maltreatment evaluations, which is the gold standard for assessing levels of concern about whether or not a child has experienced maltreatment (Berger & Lindberg, 2019). The medical evaluations are usually completed by a board-certified child abuse pediatrician or experienced advanced practice provider and a social worker, and include medical and social histories, a thorough physical exam, and a diagnostic child interview for children older than three years. These evaluations result in a medical diagnosis that includes a level of concern for maltreatment including “no”, “unlikely”, “unknown”, “suspicious”, “probable”, and “clear and confirmed”, as well as recommendations for focal child’s health and safety.

A binary variable was developed indicating children from birth to three years who (1) were evaluated for a concern of maltreatment between 3/1/2013 and 12/1/2017 at the only child abuse and neglect medical evaluation subspecialty clinic in the health system (hereafter referred to as sample evaluated for a concern of maltreatment), or (2) had at least one well-child visit in the same health system and had not been medically evaluated for a concern of maltreatment at any time between 2014 and 2017 (hereafter referred to as the general pediatric population). We did not select the first well-child visit as an index visit to avoid an oversampling of infants. Instead, one well-child visit for each child within this time frame was randomly selected as their index visit. Data before 2014 for the general pediatric population are unavailable because the health system began using a single EHR system for all practices starting from 2014 (Hurst et al., 2021).

#### Dependent Variables

##### Acute Health Service Utilization

Acute health service use was assessed by whether children had an ED visit or an overnight inpatient hospitalization 18 months following their index visit.

##### Child Maltreatment-Related Health Care Utilization

We also assessed ED visit or inpatient hospitalization possibly associated with maltreatment. To identify maltreatment-related diagnoses in our study, we applied a comprehensive list of ICD-9-CM diagnostic codes for explicit maltreatment (Supplemental Table 1) and ICD-9-CM codes suggestive of maltreatment (Supplemental Table 2) based on a previously published study (Hunter & Bernstein, 2019). Explicit maltreatment is under-documented in official health records (Hooft et al., 2015; Karatekin et al., 2018), therefore, studies have developed coding schemes using the International Classification of Diseases (ICD) codes to classify certain injuries or illnesses as suggestive of maltreatment (Lindberg et al., 2015; Schnitzer et al., 2011). Hunter and Bernstein’s (2019) study identified 62 ICD-9-CM and e-codes for suggestive maltreatment. These codes were built upon several key and seminal studies (Ben-Arieh & McDonell, 2009; King et al., 2015; Lindberg et al., 2015; Schnitzer et al., 2011). We excluded age-specific co-occurring codes (e.g., motor vehicle crash or unintentional fall) for different types of injuries to ensure that the diagnosis was associated with maltreatment rather than any other causes (Hunter & Bernstein, 2019). The presence of either a code for explicit maltreatment or suggestive maltreatment was considered to be child maltreatment-related health service use. We conducted a crosswalk from ICD-9-CM to ICD-10-CM codes to identify diagnoses related to child maltreatment using the corresponding codes for the general pediatric sample.

##### Ambulatory Care Sensitive Conditions (ACSCs)

ED visit or inpatient hospitalization for ACSCs was defined based on Lichtl et al.’s (2017) study, which identified conditions that were used and validated in at least three out of seven previously conducted studies (Anderson et al., 2012; Becker et al., 2011; Casanova et al., 1996; Flores et al., 2005; Jaeger et al., 2015; Lu & Kuo, 2012; Prezotto et al., 2015). Lichtl et al.’s (2017) study added three more conditions (allergies & allergic reactions, gastritis, and neonatal jaundice) based on local pediatricians’ opinions. We decided to use this list given these ACSCs represent a consensus from previous studies and were studied in vulnerable pediatric populations (e.g., asylum-seeking children) (Brandenberger et al., 2020; Lichtl et al., 2017). Lichtl and colleagues (2017) identified 304 ICD-10-GM (German modification) codes that were classified into 17 condition categories including 1) Allergies & allergic reactions, 2) Asthma, 3) Convulsions, 4) Dental conditions, 5) Diabetes mellitus, 6) Failure to thrive, 7) Gastritis, 8) Gastroenteritis / dehydration, 9) Immunization-preventable diseases, 10) Inflammatory diseases of female pelvic organs, 11) Iron deficiency anemia / anemia, 12) Kidney- and urinary infections, 13) Nutritional deficiency, 14) Neonatal jaundice, 15) Severe ENT & upper airway infection 16) Skin infection, and 17) Doctor’s orders have not been followed by patient. Using a framework from a prior study (Anderson et al., 2012), we assessed each of the 17 categories for inclusion to ensure relevance for a pediatric population. These considerations included (1) early access to primary care would prevent an ED visit for this condition and (2) the condition could be managed almost entirely in an ambulatory setting, assuming appropriate management (e.g., adherence to treatment). Based on this, we did not include ICD codes under the category “convulsions” given a high frequency of head trauma in our study population, which may be complicated by sequalae (e.g., subdural bleed) that lead to seizures and would require an ED visit. We created a crosswalk from ICD-10-GM to ICD-9-CM and ICD-10-CM codes for the 16 ACSCs. Our final identified ICD codes for this study are provided in Supplemental Table 3.

All dependent variables were assessed within 18 months following children’s index maltreatment evaluation visit (i.e., first visit in an episode from 2013-2017) or well-child visit (i.e., a randomly selected visit from 2014-2017). We then calculated the number of days between an index visit and the date that first outcome encounter occurred.

#### Covariates

A set of child demographic characteristics was selected for inclusion in multivariable analyses as covariates. These covariates included age (< one year of age, one year of age, two years of age, and three years of age [reference group]), sex at birth (female vs. male), and race (non-Hispanic White [reference group], non-Hispanic Black, Hispanic, and other). Health insurance was categorized by primary payor type as Medicaid and all other categories (i.e., private, other government, or others). To account for the role of socioeconomic status, we included the Area Deprivation Index (ADI) at state level in 2015, which is a ranking of neighborhood socioeconomic disadvantage. This index ranges from one to ten, with higher score indicating a higher level of disadvantage (Kind et al., 2014). We also controlled for the year of index visit (2013-2017).

### Analytic Strategy

Key characteristics of children were described using descriptive statistics including frequency, mean, standard deviation (SD), median, and interquartile range (IQR). We then conducted a descriptive bivariate analysis (i.e., test of proportions, t-test, and Mann-Whitney-Wilcoxon text) to compare health care use patterns and socio-demographic characteristics of children who were evaluated for a concern of maltreatment and the general pediatric population. For categorical variables, z-scores and 95% confidence intervals (CIs) were reported for the differences of proportions between the two samples; for continuous variable, t-score and 95% CI were reported for the differences of means between the two samples. To assess trends in time to all outcomes, we estimated Kaplan-Meier curves. Multilevel survival models were used to examine the associations between receipt of a child maltreatment evaluation and time to subsequent health care use, including first ED or inpatient visit, first maltreatment-related ED or inpatient visit, and first ED or inpatient visit for ACSCs, adjusting all pre-specified covariates. This method was chosen to account for the clustering of individual within neighborhoods. Time was defined as the number of days from an index visit to first date of an outcome, or until 18 months following an index visit (right-censored cases). We examined whether all outcome models met the proportional hazards assumptions. Findings were reported as adjusted hazard ratios (HRs), and 95% CI. P-values less than 0.05 were considered as statistically significant. Stata 16.0 was used for data management and analyses (StataCorp, 2019).

## Results

### Descriptive Analysis

Our total sample included 21,598 children. Below we describe the sample evaluated for a concern of maltreatment and the general pediatric population, respectively.

#### Description of Sample Evaluated for a Concern of Maltreatment

Descriptive characteristics of children who were evaluated for a concern of maltreatment are shown in Table 1. This sample consisted of 367 children who were evaluated at the child abuse and neglect subspecialty clinic. Among this group, about one-third (n = 107) were under one year of age. The children were predominantly non-Hispanic Black (n = 213, 58.0%) and nearly 80% (n = 288) of this sample were covered by Medicaid. The median ADI score was 5 (IQR = 5.0). Notably, about 43.3% (n = 159) of children had ED visits or inpatient hospitalizations in 18 months following an index evaluation and 3.3% (n = 12) had visits related to maltreatment. Of the children who had a subsequent ED or inpatient visit, 7.6% (12/159) of those visits were explicit for or suggestive of maltreatment. Around 15.3% (n = 56) of children had visits for ACSCs.

**Table 1.** Descriptive statistics of sample evaluated for a concern of maltreatment and general pediatric population (N = 21,598)

| Characteristics | Sample evaluated<br>for maltreatment,<br>n (%)<br>(n = 367) | General pediatric<br>population,<br>n (%)<br>(n = 21,231) | Test<br>results | 95% Confidence<br>Interval |
| --- | --- | --- | --- | --- |
| Emergency department<br>use/inpatient<br>hospitalization | 159 (43.3) | 6415 (30.2) | 5.4 | 8.0, 18.2*** |
| Maltreatment-related<br>emergency department<br>use/inpatient<br>hospitalization | 12 (3.3) | 133 (0.6) | 6.2 | 0.8, 4.5*** |
| Emergency department<br>use/inpatient<br>hospitalization for<br>ambulatory care sensitive<br>conditions | 56 (15.3) | 2259 (10.6) | 2.8 | 0.9, 8.3** |
| Age at first encounter |  |  |  |  |
| < 1 year | 107 (29.2) | 10015 (47.2) | -6.9 | -22.7, -13.3*** |
| 1 year | 76 (20.7) | 4527 (21.3) | -0.3 | -4.8, 3.6 |
| 2 years | 84 (22.9) | 2819 (13.3) | 5.4 | 5.3, 13.9*** |
| 3 years | 100 (27.3) | 3870 (18.2) | 4.4 | 4.4, 13.6*** |
| Race/ethnicity |  |  |  |  |
| Black, Non-Hispanic | 213 (58.0) | 7258 (34.2) | 9.5 | 18.8, 28.9*** |
| White, Non-Hispanic | 64 (17.4) | 6849 (32.3) | -6.0 | -18.8, -10.9*** |
| Hispanic | 59 (16.1) | 3586 (16.9) | -0.4 | -4.6, 3.0 |
| Other or Unknown | 31 (8.5) | 2538 (16.7) | -4.2 | -11.1, -5.3*** |
| Sex |  |  |  |  |
| Female | 197 (53.7) | 10406 (49.0) | 1.8 | -0.5, 9.8 |
| Male | 170 (46.3) | 10825 (51.0) | -1.8 | -9.8, 0.5 |
| Health insurance status |  |  |  |  |
| Medicaid | 288 (78.5) | 9788 (46.1) | 12.3 | 28.1, 36.6*** |
| All other types | 79 (21.5) | 11443 (53.9) | -12.3 | -36.6, -28.1*** |
| Area Deprivation Index,<br>mean (SD) | 5.3 (2.8) | 4.1 (2.6) | 8.0 | 0.9, 1.5*** |
| Area Deprivation Index,<br>median (IQR) | 5.0 (5.0) | 3.0 (4.0) | 8.2 | 0.2, 0.3*** |

*Note.* SD, standard deviation; IQR, interquartile range; percentages of categorical variables were compared using test of proportions and the z-scores were reported; means of Area Deprivation Index were compared using t-test and the t-score was reported; medians of Area Deprivation Index were compared using Mann-Whitney-Wilcoxon test and the z-score was reported. Z-scores, t-scores, and 95% confidence intervals were reported for the differences of proportions or means between the two samples. \*Significant at the $p$ -value < 0.05 level; \*\*significant at the $p$ -value < 0.01 level; \*\*\*significant at the $p$ -value < 0.001 level.

#### Description of the General Pediatric Population

The general pediatric population who were not evaluated for a concern of maltreatment at the subspecialty clinic consists of 21,231 children (Table 1). Among this sample, almost 50% (n = 10,015) were under one year old. The distribution of race/ethnicity was approximately equal between non-Hispanic White (n = 6,849, 32.3%) and non-Hispanic Black (n = 7,258, 34.2%), and slightly less than half of children were covered by Medicaid (n = 9,788, 46.1%). This group of children had a median ADI score of 3 (IQR = 4.0). Approximately 30.2% (n = 6,415) had a subsequent ED visit or inpatient hospitalization and 0.6% (n = 133) had maltreatment-related ED visits or inpatient hospitalizations. Thus, 2.1% (133/6415) of the ED or inpatient visits were explicit for or suggestive of maltreatment. About 10.6% (n = 2,259) of children presented for ED or inpatient care for ACSCs.

### Bivariate Analysis

Table 1 shows results for bivariate comparisons between the sample evaluated for a concern of maltreatment and the general pediatric population. The tests of proportions indicated that compared to the general pediatric population, children evaluated for a concern of maltreatment were significantly more likely to have subsequent acute health care use, including ED visits or inpatient hospitalizations in general (43.3% vs. 30.2%, *z* = 5.4, 95% CI [8.0, 18.2]), maltreatment-related ED visits or inpatient hospitalization (3.3 % vs. 0.6%, *z* = 6.2, 95% CI [0.8, 4.5]), and ED visits or inpatient hospitalization for ACSCs (15.3% vs. 10.6%, *z* = 2.8, 95% CI [0.9, 8.3]) in 18 months following an index visit. Kaplan-Meier curves were demonstrated in Figure 1, indicating that the probabilities of different acute health care use patterns were constantly higher among children who were evaluated for a concern of maltreatment as compared to the general pediatric population.

**Figure 1.**
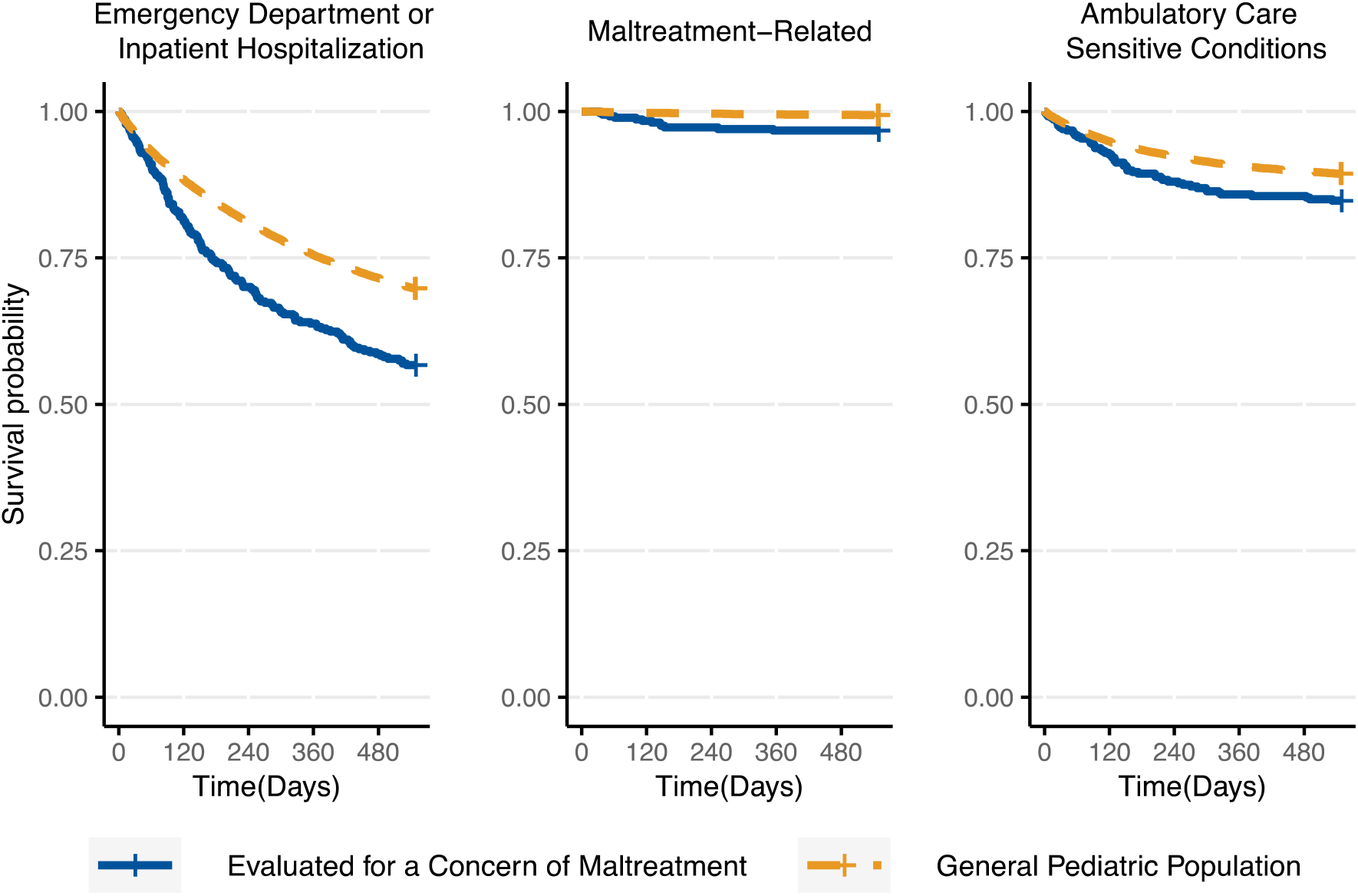
Kaplan-Meier curves describing acute health care use within 18 months after index encounter among children birth to age three years

Children evaluated for a concern of maltreatment were less likely to be younger than one year of age (29.2% vs. 47.2%, *z* = -6.9, 95% CI [-22.7, -13.3]), and more likely to be non-Hispanic Black (58.0% vs. 34.2%, *z* = 9.5, 95% CI [18.8, 28.9]), Medicaid-enrolled (78.5% vs. 46.1%, *z* = 12.3, 95% CI [28.1, 36.6]), and live in more disadvantaged neighborhoods (mean of ADI = 5.3 vs. 4.1, *t =* 8.0, 95% CI [0.9, 1.5]) than the general pediatric population.

### Multivariable Analysis

All of our multivariate models met the proportional hazards assumptions. Overall, models were significant for the associations between a receipt of maltreatment evaluation and an acute health care use (Wald χ^2^ = 2024.4, *df* = 14, *p* < 0.001), child maltreatment related acute health care use (Wald χ^2^ = 125.9, *df* = 14, *p* < 0.001), and acute health care use for ACSCs (Wald χ^2^ = 1056.3, *df* = 14, *p* < 0.001). Results of the multivariable analysis (Table 2) indicated that after adjusting for child age, sex, race/ethnicity, health insurance status, and ADI, children who were evaluated for a concern of maltreatment had an increased hazard ratio for any type of subsequent ED visit or inpatient hospitalization (HR: 1.3, 95% CI [1.1, 1.5]), and a subsequent maltreatment-related visit (HR: 4.4, 95% CI [2.3, 8.2]) relative to the general pediatric population. However, receipt of an evaluation for maltreatment was not associated with higher hazard of a subsequent health care use for ACSCs (HR: 1.0, 95% CI [0.7, 1.3]).

**Table 2.** Cox regression results: Association between receipt of a maltreatment medical evaluation and time until acute health care use with 95% confidence intervals (N = 21,598)

| Cox Model | Any emergency department or inpatient visit |  | Any maltreatment-related emergency department or inpatient visit |  | Any emergency department or inpatient visit for ACSCs |  |
| --- | --- | --- | --- | --- | --- | --- |
|  | HR | 95% CI | HR | 95% CI | HR | 95% CI |
| Children received medical evaluation for a concern of maltreatment | 1.3 | 1.1, 1.5** | 4.4 | 2.3, 8.2*** | 1.0 | 0.7, 1.3 |
| Age at first encounter |  |  |  |  |  |  |
| < 1 year | 2.1 | 1.9, 2.2*** | 1.7 | 1.0, 2.9 | 2.3 | 2.0, 2.6*** |
| 1 year | 1.6 | 1.5, 1.8*** | 1.7 | 0.9, 3.0 | 1.7 | 1.5, 2.0*** |
| 2 years | 1.2 | 1.1, 1.4*** | 1.3 | 0.7, 2.5 | 1.3 | 1.1, 1.5** |
| 3 years (ref) |  |  |  |  |  |  |
| Race/ethnicity |  |  |  |  |  |  |
| White, Non-Hispanic (ref) |  |  |  |  |  |  |
| Black, Non-Hispanic | 1.9 | 1.8, 2.1*** | 2.1 | 1.2, 3.6*** | 2.9 | 2.5, 3.3*** |
| Hispanic | 1.7 | 1.6, 1.9*** | 0.9 | 0.5, 1.8 | 2.2 | 1.9, 2.6*** |
| Other or Unknown | 1.1 | 1.0, 1.2 | 0.7 | 0.3, 1.5 | 1.3 | 1.1, 1.5** |
| Sex |  |  |  |  |  |  |
| Female | 0.9 | 0.8, 0.9*** | 0.9 | 0.7, 1.3 | 0.9 | 0.8, 0.9** |
| Male (ref) |  |  |  |  |  |  |
| Health insurance status |  |  |  |  |  |  |
| Medicaid | 1.8 | 1.7, 1.9*** | 2.9 | 1.8, 4.5*** | 2.2 | 1.9, 2.4*** |
| All other types (ref) |  |  |  |  |  |  |
| Area Deprivation Index | 1.1 | 1.1, 1.1*** | 1.0 | 1.0, 1.1 | 1.1 | 1.0, 1.1*** |
| Model Fit | Wald $\chi^2 = 2024.4$, $df = 14$, $p < 0.001$ | | Wald $\chi^2 = 125.9$, $df = 14$, $p < 0.001$ | | Wald $\chi^2 = 1056.3$, $df = 14$, $p < 0.001$ | |

*Note.* Abbreviations: ACSC, ambulatory care sensitive condition; HR, hazard ratio; CI, confidence interval; ref, reference group for categorical variables; *df*, degree of freedom. Controlling for year of index child maltreatment visit. \*Significant at the $p$ -value $< 0.05$ level; \*\*significant at the $p$ -value $< 0.01$ level; \*\*\*significant at the $p$ -value $< 0.001$ level.

## Discussion

Our study aimed to assess whether a child maltreatment evaluation was associated with a higher risk of subsequent health care use over an 18-month period. Our findings revealed that, compared to the general pediatric population who were not evaluated for maltreatment, receiving a medical evaluation for a concern of maltreatment was significantly associated with higher hazards of subsequent ED visits or inpatient hospitalizations and maltreatment-related visits. The observed associations could not be explained by differences in age, sex, race/ethnicity, or health insurance status.

The prevalence of an ED visit or inpatient hospitalization for children who were evaluated for a concern of maltreatment was much higher than that in the general pediatric population in our study, as well as previous estimates for general pediatric samples (34%-38%) (Lee & Monuteaux, 2019; McDermott et al., 2018). These significant differences still existed even when we accounted for race/ethnicity and Medicaid enrollment, two salient factors affecting health care use (Hong et al., 2007; Riera & Walker, 2010).

We found that children who were evaluated for a concern of maltreatment were approximately four times as likely as the general pediatric population to visit ED or inpatient care for maltreatment or for injuries, illnesses, or conditions associated with maltreatment. The proportions of maltreatment-related visits in all ED or inpatient visits between the evaluated children and the general pediatric population (7.6% vs 2.1%) is similar to proportions found in prior studies that examined this in general pediatric populations (Hunter & Bernstein, 2019; King et al., 2015). This substantial difference suggests that even after services and referrals were provided, children evaluated for a concern of maltreatment were still at a higher risk of future harm or complications from maltreatment evaluated at the index visit.

Children who were evaluated for a concern of maltreatment had a higher rate of visiting an ED or inpatient care for ACSCs than the general pediatric population. Yet when we controlled for socio-demographic characteristics that might confound this association in multivariate analyses, an evaluation for maltreatment was not associated with a higher probability of health care visits for ACSCs relative to the general pediatric population. Despite this, our study confirmed several previously identified demographic and socioeconomic factors that were potentially associated with preventable ED use or hospitalization (Bettenhausen et al., 2017; Kangovi et al., 2013). As previous studies have shown, children at risk for maltreatment are more likely to reside in areas with high levels of poverty and social, political, and economic disenfranchisement, and where systemic resources to support parents and families are limited (Kim & Drake, 2018). Such living environment and socioeconomic status (SES) may lead to elevated risk for hospitalizations for ACSCs (Bettenhausen et al., 2017; Kangovi et al., 2013; Wallar et al., 2020) because children from low SES families are more likely to be injured than those from high SES backgrounds and use acute health services as their primary source of medical care than ambulatory care.

Although the nonsignificant association between a maltreatment evaluation and increased use of ED or inpatient care for ACSCs was unexpected, it reflects a more important role of non-dominant racial/ethnic groups, Medicaid enrollment, and low SES in driving a high rate of accessing acute health care for ACSCs. This is possibly caused by poor parental health literacy (Sanders et al., 2009), restricted access to transportation, limited availability of primary care in this population, or perceptions that hospitals offer better access, health care quality, and technical competence (Kangovi et al., 2013). Due to limited studies in this area, future pediatric research should explore measurements of ACSCs and better understand its relationship with children’s use of health care and potential complex relationships with SES and race/ethnicity.

This study indicates that it is important to characterize health care utilization by the nature of services, which allows for targeted interventions for children with various utilization patterns. In addition to maltreatment related health encounters and ACSCs, there are still a large proportion of children who have experienced maltreatment or have had a concern for maltreatment present to ED or inpatient care with other complaints or diagnoses, though not necessarily ACSCs. We did not assess chief complaint or diagnosis that could provide an explanation for the high rate of utilization. Previous literature is limited regarding diagnostic differences between maltreated and general pediatric patients. Some studies suggested that although compared to population-based control subjects, children with confirmed child abuse had almost twice risk of ED visits before their visits for maltreatment symptoms, these two groups were similar in symptoms presented in ED visits (Bailhache et al., 2022; Guenther et al., 2009). Since child maltreatment is associated with a variety of negative behavioral, emotional, and health outcomes leading to ED or hospitalization (Jonson-Reid et al., 2012), our study reinforces a need for investigating other potential reasons or utilization patterns that may lead to the high rate of acute health care use among high-risk cohorts.

### Limitations

There were several notable limitations with our study. Our data were from one academic medical system and, therefore, our findings may not be more broadly representative. While sufficient for addressing our objectives, a relatively small sample of children who have been evaluated for a concern of maltreatment prevented us from exploring specific types of maltreatment. Second, although our data source is reliable, ICD codes for maltreatment are often documented as under-utilized (Scott et al., 2009). This could potentially result in an underestimation of maltreatment-related health care use. However, we used a solid algorithm to identify suspected maltreatment in EHR, which helped more accurately quantify this public health problem. Finally, data on index dates for children evaluated for a concern of maltreatment and the general pediatric population were not matched. A study period of 2014-2019 was chosen for the general pediatric population given data availability over this time. Our results may be subject to immortal time bias (Lévesque et al., 2010), and the interpretation of HRs should be conservative. Data are also not available for children who were censored due to competing risk events (Satagopan et al., 2004), such as dying, moving out of the county, or other causes. However, we assumed that only a small proportion of our sample have experienced such events, thus our findings are unlikely to have been affected dramatically by not accounting for competing risks.

### Implications for Practice

Our study has clinical and research implications regarding early prevention and ambulatory care use of children who have been evaluated for a concern of maltreatment. Our findings highlight that these children had a higher risk of acute health service use, both for maltreatment-related injuries or illnesses and conditions more broadly, even after potential maltreatment has been diagnosed, involvement with child welfare when mandated, and when other referrals and supports were offered. Characterizing health care utilization by patterns of service use would allow for targeted preventative measures and anticipatory guidance to help families and children with different medical needs seek care from appropriate general or subspecialty pediatricians. Clinicians in the child abuse and neglect subspecialty clinic could provide appropriate health service recommendations to high-risk children and families, and work with community partners to ensure children’s adherence to the recommended or placed services. Clinicians could ensure children who have been evaluated for a concern of maltreatment have access to routine care with a trusted healthcare provider. This provider can continue to monitor children’s health and proactively provide advice and guidance should there be complications of existing conditions. Furthermore, to minimize risks of escalating or repeated maltreatment, healthcare providers in hospitals and EDs should be aware of children’s maltreatment history and target children who have maltreatment-related diagnoses as a high-risk group to prevent ongoing or further victimization. Given that many children who have been evaluated for a concern of maltreatment used health services not specific to maltreatment, clinicians should also pay attention to other health needs separate from concerns of immediate harm.

To avoid potentially preventable ED or hospital utilization, reduce cost of care, and improve patient experience, access to appropriate ambulatory care should be improved for non-Hispanic Black and Hispanic patients, and low-SES patients. It should be noted that very few studies have assessed ACSCs for children who have possibly experienced maltreatment, and this issue needs to be investigated further in other samples (e.g., children involved in child protective services). Relative to the general pediatric population, children with suspected maltreatment have more chronic illnesses and higher medical complexity (Azzopardi et al., 2021), and therefore may have certain reasons to make health care use decisions. To identify potentially modifiable factors and develop appropriate interventions, more in-depth studies are needed on parent perspectives about the reasons that drive their use of ambulatory care for children at risk of maltreatment.

## Conclusion

This study examined the association between receiving a maltreatment evaluation and risk of subsequent health care use among children from birth to age three. Specifically, we found that a receipt of maltreatment evaluation was associated with higher risk for a subsequent ED visit or inpatient hospitalization, and a maltreatment-related visit. Hospitals and EDs should pay attention to complications of maltreatment, maltreatment history, and health needs of high-risk children. Child abuse and neglect clinicians could utilize these patterns of health care use to identify concerning trends and help high-risk families minimize complications resulting from previous maltreatment or prevent future harm.

## Data Availability

Data not available due to ethical restrictions.

