## Supplemental Table 1 for "Do children evaluated for maltreatment have higher subsequent emergency department and inpatient care utilization compared to a general pediatric sample?"

Supplemental Table 1. International Classification of Diseases, Ninth Revision, Clinical Modification (ICD-9-CM) codes specific for child maltreatment

| Code Description | ICD-9-CM Codes |
| --- | --- |
| Child maltreatment syndrome | 995.5 |
| Child abuse, unspecified | 995.50 |
| Child emotional/psychological abuse | 995.51 |
| Child neglect (nutritional) | 995.52 |
| Child sexual abuse | 995.53 |
| Child physical abuse | 995.54 |
| Shaken baby syndrome | 995.55 |
| Other child abuse and neglect | 995.59 |
| Perpetrator of child and adult abuse |  |
| father, stepfather, or boyfriend | E967.0 |
| other specified person | E967.1 |
| mother, stepmother, or girlfriend | E967.2 |
| sibling | E967.5 |
| grandparent | E967.6 |
| other relative | E967.7 |
| non-related caregiver | E967.8 |
| unspecified person | E967.9 |
