## Supplemental Table 2 for "Do children evaluated for maltreatment have higher subsequent emergency department and inpatient care utilization compared to a general pediatric sample?"

Supplemental Table 2. International Classification of Diseases, Ninth Revision, Clinical Modification (ICD-9-CM) codes for suggestive child maltreatment

| Code Description | ICD-9-CM Codes | Age Included (years) |
| --- | --- | --- |
| **Suggestive physical maltreatment** |  |  |
| Observation for abuse/neglect | V71.81 | <10 |
| Retinal hemorrhage^d,e^ | 362.81 | <3 |
| Rib fracture^d,e^ | 807.0; 807.1 | <5 |
| Scapula fracture^d,e^ | 811 | <5 |
| Other/unspecified intracranial hemorrhage^d,e^ | 853 | <5 |
| Stomach injury^d,e^ | 863.1 | <10 |
| Assault | E965; E966; E968.2 | <4 |
| Assault, NOS | E968.9 | <4 |
| Undetermined intent, other means | E988 | <10 |
| Femur fracture^d,e^ | 820; 821 | <1 |
| Humerus fracture^d,e^ | 812 | <1 |
| **Suggestive neglect and physical maltreatment ^d,e^** |  |  |
| Skull vault fracture | 800 | <5 |
| Vertebral fracture | 805 | <5 |
| Traumatic subarachnoid hemorrhage | 852 | <5 |
| Intrathoracic injury, NEC | 862 | <5 |
| Spleen injury | 865 | <5 |
| Spinal cord injury | 952 | <3 |
| **Suggestive neglect** |  |  |
| Other severe malnutrition | 262 | <10 |
| Dental carries | 521 | <10 |
| Solar radiation dermatitis | 692.7 | <2 |
| Pelvic fracture^d,e^ | 808 | <5 |
| Heart or lung injury^d,e^ | 861 | <5 |
| Liver injury^d,e^ | 864 | <5 |
| Kidney injury^d,e^ | 866 | <5 |
| Burn of head^f^ | 941 | <5 |
| Burn of trunk^f^ | 942 | <5 |
| Burn of multiple sites^f^ | 946 | <5 |
| Poisoning by drugs, medicinal^g^ | 960-979 | <5 |
| Second–hand tobacco smoke | E869.4 | <10 |
| Drowning, nonfatal submersion | 994.1 | <4 |
| Bathtub (near) drowning | E910.4 | <4 |
| Other (near) drowning | E910.8 | <4 |
| Accidental (near) drowning, NEC | E910.9 | <4 |
| Unarmed fight, brawl | E960.0 | <4 |
| **Suggestive sexual maltreatment** |  |  |
| Genital herpes^a^ | 54.1 | <10 |
| Gonococcal infection^b^ | 98.0 | <10 |
| Pelvic inflammatory disease, unspecified | 614.9 | <10 |
| Contusion of genital organs^c,d,e^ | 922.4 | <10 |
| Observation after alleged rape | V71.5 | <10 |

*Notes.* Exclusion of co-occurring codes:

a. Other congenital infections specific to perinatal period (771.2).

b. Gonococcal infection of eye neonatal conjunctivitis (98.4, 771.6).

c. Coagulation defects (286, 287).

d. Motor vehicle crash (E800-E819).

e. Unintentional fall (E880-E888).

f. Burns from house or structural fire (E890-E899).

g. Medical misadventures (E876).
