## Supplemental Table 3 for "Do children evaluated for maltreatment have higher subsequent emergency department and inpatient care utilization compared to a general pediatric sample?"

Supplemental Table 3. International Classification of Diseases, Ninth Revision, Clinical Modification (ICD-9-CM) codes for ambulatory care sensitive conditions

| Pediatric Ambulatory Care Sensitive Conditions | ICD-9-CM Codes |
| --- | --- |
| Allergies & allergic reactions | 477.0, 477.1, 477.2, 477.8 477.9, 558.3, 692.83, 692.4, 692.81, 692.3, 692.89, 692.4, 692.5, 692.6, 692.84, 692.89, 692.9, 995.3, V58.89, 909.9, 995.21, V14 |
| Asthma | 493 |
| Dental | 521, 522, 523, 525, 528 |
| Diabetes | 250.0, 250.1, 250.2, 250.3, 250.8, 250.9 |
| Doctor’s orders have not been followed by the patient | V45.12, V15.81 |
| Ear, nose and throat (ENT) | 382, 462, 463, 465, 472.1 |
| Failure to thrive | 783.3, 783.4 |
| Gastritis | 535 |
| Gastroenteritis/dehydration | 008.6, 008.8, 009, 558.9, 276.5 |
| Immunization-preventable conditions | 032, 033, 037, 045, 052, 055, 056, 070.2, 070.3, 072, 320.0, 390, 391 |
| Iron-deficiency anemia | 280.1, 280.8, 280.9 |
| Kidney and urinary infections | 590, 599.0, 599.9 |
| Neonatal jaundice | 774.3, 774.5, 774.6 |
| Nutritional deficiencies | 260, 261, 262, 268.0, 268.1 |
| Pelvic inflammatory disease | 614 |
| Skin infection | 681.0, 681.1, 682.0, 682.2, 682.3, 682.4, 682.5, 682.6, 682.07, 682.8, 682.9 |
